# Assessing the impact of adherence to Non-pharmaceutical interventions and indirect transmission on the dynamics of COVID-19: a mathematical modelling study

**DOI:** 10.1101/2021.08.16.21262135

**Authors:** Sarafa A. Iyaniwura, Musa Rabiu, Jummy F. David, Jude D. Kong

**Affiliations:** Department of Mathematics and Institute of Applied Mathematics, University of British Columbia, Vancouver, BC, Canada; School of Mathematics, Statistics & Computer Science, University of KwaZulu-Natal, Durban, South Africa; Department of Mathematics and Statistics, York University, Toronto, Ontario, Canada; Canadian Centre for Diseases Modeling (CCDM), York University, Toronto, Ontario, Canada; Laboratory of Mathematical Parallel Systems (LAMPS), York University, Toronto, Ontario, Canada

**Keywords:** COVID-19, epidemics, SARS-CoV-2, direct and indirect transmission, seir models, non-pharmaceutical intervention, adherent and non-adherent, population dynamics

## Abstract

Adherence to public health policies such as the non-pharmaceutical interventions implemented against COVID-19 plays a major role in reducing infections and controlling the spread of the diseases. In addition, understanding the transmission dynamics of the disease is also important in order to make and implement efficient public health policies. In this paper, we developed an SEIR-type compartmental model to assess the impact of adherence to COVID-19 non-pharmaceutical interventions and indirect transmission on the dynamics of the disease. Our model considers both direct and indirect transmission routes and stratifies the population into two groups: those that adhere to COVID-19 non-pharmaceutical interventions (NPIs) and those that do not adhere to the NPIs. We compute the control reproduction number and the final epidemic size relation for our model and study the effect of different parameters of the model on these quantities. Our results show that direct transmission has more effect on the reproduction number and final epidemic size, relative to indirect transmission. In addition, we showed that there is a significant benefit in adhering to the COVID-19 NPIs.

## 1 Introduction

The severe acute respiratory syndrome coronavirus 2 (SARS-CoV-2) was first discovered in the city of Wuhan, Hubei province, China, in December 2019 [54] and has since spread all over the world with over 181.1 million reported cases and over 3.9 million deaths globally as of June 29, 2021 [53]. Following the outbreak of COVID-19, several non-pharmaceutical interventions (NPIs) such as physical distancing, isolation, hand washing, stay-at-home order, closing of schools and businesses, travel restrictions, among others, were implemented all over the world to limit the spread of the disease [24, 25, 33, 45]. Despite implementing these NPIs, there is still a significant number of COVID-19 cases and deaths reported daily. On January 20, 2020, the World Health Organization (WHO) declared COVID-19 a public health emergency of international concern (PHEIC) [57] and a pandemic on March 11, 2020 [58].

Apart from the increasing number of cases and deaths, the COVID-19 pandemic has had a significant effect on our day-to-day activities such as socializing, entertainment, education, tourism, health, and businesses. Since the outbreak of the SARS-CoV-2 virus, researchers have been studying its transmission dynamics. The virus is believed to be transmitted through two major pathways namely: the direct and indirect pathways [30, 15, 44, 55, 26]. The direct pathway, also known as human-to-human transmission, includes transmission through inhalation of virus-infected droplets, coughing, sneezing and having physical contact with infected persons. Indirect transmission may occur when a susceptible individual comes in contact with a contaminated commonly shared surface or object. These contamination usually occur when an infected individual touches, coughs or sneezes on the objects.

Several mathematical models have been developed to study the transmission dynamics of COVID-19 [32, 23, 28, 63, 2, 43, 50, 13, 3, 38, 42]. In [38], two differential equation models were developed to study the effect of the exposed or latency period on the dynamics of COVID-19 in China. Another model was developed in [42] to study the effect of super-spreaders on the dynamics of COVID-19 in Wuhan, China. These and many other mathematical models have provided a lot of insights into the dynamics of COVID-19. During the early stage of the COVID-19 pandemic, physical distancing, wearing of face masks, hand washing, and other non-pharmaceutical interventions (NPIs) were the primary interventions against the disease worldwide. Many mathematical models have been used to study the impact of these NPIs on the dynamics of COVID-19. This includes the susceptible-exposed-infected-quarantine-recovered (SEIQR) model developed in [3] to study the impact of physical distancing on the dynamics of COVID-19 in British Columbia (BC), Canada, and the model of [50] used to study the impact of different mitigation strategies on the dynamics of COVID-19 in Ontario, Canada. In [5], a susceptible–exposed–infected–removed (SEIR) model was designed on a temporal network, which evolves according to the activity-driven paradigm. Their model was used to study the spread of the disease as a function of the fraction of the population following public health measures. Their result shows that physical distancing and mask-wearing can effectively prevent COVID-19 outbreaks if adherence to both measures involves a substantial fraction of the susceptible population. The effect of timing of adherence to COVID-19 NPIs such as social distancing on COVID-19 in the United States was studied using an agent-based model in [1]. Their model was applied to Dane County, Wisconsin, the Milwaukee metropolitan area and New York City (NYC), and show that the timing of implementing and easing the social distancing measures has major effects on the number of COVID-19 cases. The results obtained in the works of [49, 52] are also in the same line with the findings mentioned above. Many of these models focus on direct transmission of COVID-19 through in-person contacts despite the emphasis by the world health organization (WHO) on indirect transmission of the disease [55].

A few researchers have studied indirect transmission of COVID-19 [63, 2, 43, 19, 41, 40]. In [19], a novel deterministic susceptible-exposed-infected-removed-virus-death (SEIRVD) model was developed to study the potential impact of both direct and indirect transmission of COVID-19 on the dynamics of the disease in Ontario, Canada. Their results showed significant increase in number of cases with indirect transmission. They also highlighted the importance of implementing additional preventive and control measures to minimize the spread of COVID-19 through both transmission routes. A five-compartment model for both direct and indirect transmission pathways was developed in [41], and used to study the impact of both transmission pathways on the dynamics of COVID-19. The mathematical model of [40] also used indirect transmission mechanisms to study the dynamics of COVID-19. They examined the conditions under which contaminated objects may lead to a significant spread of SARS-CoV-2 during and after lock-down using a SEIR model with the addition of a fomite term. They proposed that the addition of a fomite term will help to better understand the transmission dynamics of the virus and also in policy making. To the best of our knowledge, none of these studies have looked at the effect of adherence and non-adherence to COVID-19 NPIs on both direct and indirect transmissions of the disease.

In this study, we develop an SEIRV compartmental model for studying the transmission dynamics of COVID-19. Our model considers both direct and indirect transmission of the disease following the approach of [19]. It has a similar structure to the model of [3] used to study the impact of physical distancing on the contact rates in British Columbia (BC), Canada, where BC population was divided into the groups practicing physical distancing and those not practicing physical distancing. Here, we stratify our population into two groups: those that adhere to all COVID-19 non-pharmaceutical interventions (NPIs) and those that do not adhere. We compute the control reproduction number and final epidemic size for our model, and study the effect of different parameters of the model on these quantities. This model is used to assess the impact of adherence to COVID-19 non-pharmaceutical interventions and indirect transmissions on the dynamics of the disease. Our study highlights the importance of adhering to the COVID-19 NPIs as instructed by the World Health Organization (WHO) in reducing infections and eventually controlling the spread of the disease.

The remaining part of this paper is organized as follows: The model is developed in Section 2. In Section 3, the control reproduction number of the disease is calculated using the next generation matrix approach. We also computed a final size relation for the epidemic in this section. In Section 4, we numerically study the effect of important model parameters on the control reproduction number calculated in Section 3. In addition, we present numerical simulations of the SEIRV model for different scenarios in this section. These simulations are used to comprehend the effect of direct and indirect transmission, with or without adherence on the dynamics of the disease. We conclude the paper in Section 5 with a brief discussion.

## 2 Mathematical model

A compartmental SEIRV model of COVID-19 epidemic is developed and analyzed. The model consider both direct and indirect transmission routes and divides the population into two main groups: adherent and non-adherent populations. The adherent population include individuals in the population that adhere to all COVID-19 non-pharmaceutical interventions (NPIs), while the non-adherent population contains those that do not adhere to the NPIs. The compartments of the model for non-adherent population are defined as follows: susceptible (*S*); exposed (*E*); infected (*I*); and recovered/removed (*R*). Corresponding compartments for the adherent population are given by *S*_*a*_, *E*_*a*_, *I*_*a*_, and *R*_*a*_, respectively. Note that individuals in the exposed compartments *E* and *E*_*a*_ are not infectious. We assume that recovered individuals have permanent immunity from the disease (no reinfection). The compartment *V* accounts for the virus shed on contaminated surfaces and objects by infected individuals. Figure 1 shows a schematic diagram of the model, where the solid black arrows indicate the direction of the flow of individuals between the compartments at the rates indicated beside the arrows. The dashed red arrows show virus shedding or contamination of surfaces by infected individuals. The differential equations for non-adherent population are given by

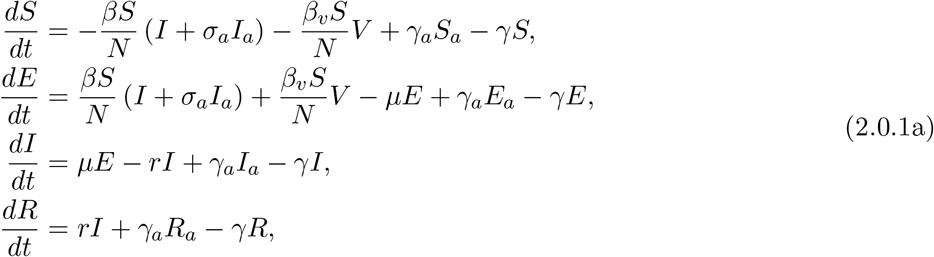

where *β* and *β*_*v*_ are the direct and indirect transmission rates, respectively, for non-adherent population. The parameter *σ*_*a*_, with 0 ≤ *σ*_*a*_ ≤ 1 is used to model the reduction in virus shedding (or contamination of surfaces), susceptibility, and onward transmission of disease due to adherence to NPIs. Parameters *γ*_*a*_ and *γ* are the rates of movement from adherence to non-adherence and vice versa, respectively, while *µ* denotes the rate of transitioning from the exposed compartment *E* to the infected compartment *I*, and *r* is the recovery rate. The corresponding equations for the adherent population are given by

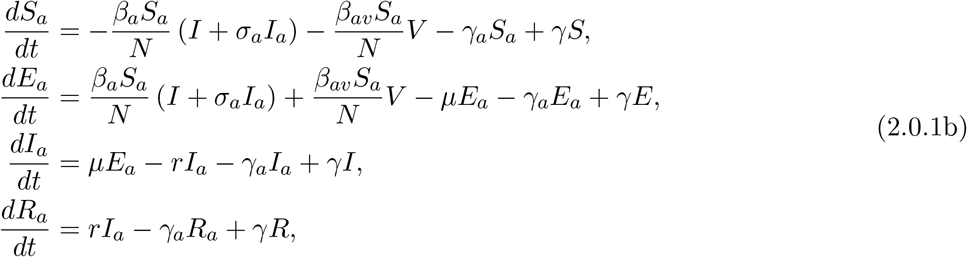

where *S*_*a*_, *E*_*a*_, *I*_*a*_ and *R*_*a*_ are for susceptible, exposed, infected, and recovered individuals in the adherent population, respectively. The parameters *β*_*a*_ and *β*_*av*_ are the direct and indirect disease transmission rates, respectively, for the adherent population. We assume that individuals in both adherent and non-adherent population transition from exposed to infected compartments, and recover from the disease at the same rates, *µ* and *r*, respectively. In addition, we assume that individuals move from adherent to non-adherent and vice versa, at the same rates in the susceptible, exposed, infected, and recovered compartments. The disease dynamics for both non-adherent (2.0.1a) and adherent (2.0.1b) populations are coupled to the dynamics of the virus in the environment or on contaminated surfaces given by

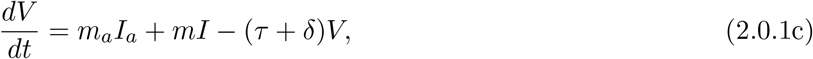

where *m* and *m*_*a*_ are the virus shedding (or surface contamination) rates for non-adherent and adherent populations, respectively. The parameter *δ* is the decay rate of SAR-CoV-2 in the environment and on surfaces, and *τ* is the environment cleaning/sanitization rate. We assume that our population is constant throughout the epidemic period so that *N* = *N*_⋆_(*t*) + *N*_*a*_(*t*) for all time *t* ≥ 0, where *N* (*t*) = *S*(*t*) + *E*(*t*) + *I*(*t*) + *R*(*t*) and *N*_*a*_(*t*) = *S*_*a*_(*t*) + *E*_*a*_(*t*) + *I*_*a*_(*t*) + *R*_*a*_(*t*). In our SEIRV model (2.0.1), the compartments *S, E, I, R, S*_*a*_, *E*_*a*_, *I*_*a*_, *R*_*a*_ have dimension of *individual*, while the virus compartment *V* has dimension of *virus particle*. The dimensions of the model parameters are given as follows:

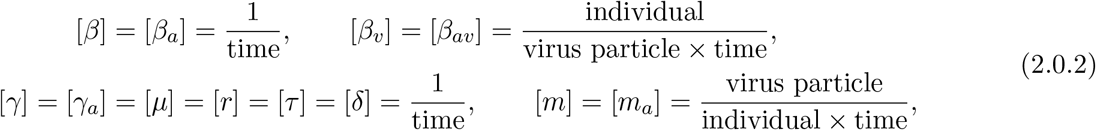

where [*λ*] represents dimension of *λ*. Note that *σ*_*a*_ is a dimensionless parameter in our model. It is important to mention that there are relationships between the parameters *β, β*_*v*_, and *m* for non-adherent population and *β*_*a*_, *β*_*av*_, and *m*_*a*_ of adherent population, respectively. These relationships are used when computing the control reproduction number in Section 3.1 and final size relation in Section 3.2. We define the fraction of adherence in the population as *f* = *γ/*(*γ* + *γ*_*a*_), on which the dynamics of the ODE system (2.0.1) depends. Since adherence to COVID-19 NPIs reduces transmission rate for the adherent population, we equate the direct and indirect transmission rates for the adherent population to those of non-adherent population as follow

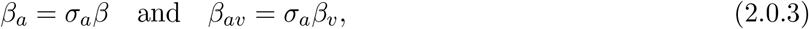

where 0 ≤ *σ*_*a*_ ≤ 1. When *σ*_*a*_ = 0, we have *β*_*a*_ = 0, *β*_*av*_ = 0 and *σ*_*a*_*I*_*a*_ = 0 in (2.0.1a) and (2.0.1b), which implies that adherence to NPIs is 100% effective in preventing infections in the adherent population, and that adherent individuals do not infect the non-adherent individuals. On the other hand, when *σ*_*a*_ = 1, we have *β*_*a*_ = *β, β*_*av*_ = *β*_*v*_ and *σ*_*a*_*I*_*a*_ = *I*_*a*_, which implies that disease is transmitted at the same rate in both adherent and non-adherent populations. Table 1 shows the model variables and their description.

**Table 1:** Model variables and description.

| Variable | Description |
| --- | --- |
| $S$ | Non-adherent susceptible population |
| $E$ | Non-adherent exposed population |
| $I$ | Non-adherent infected population |
| $R$ | Non-adherent recovered population |
| $S_a$ | Adherent susceptible population |
| $E_a$ | Adherent exposed population |
| $I_a$ | Adherent infected population |
| $R_a$ | Adherent recovered population |

**Figure 1:**
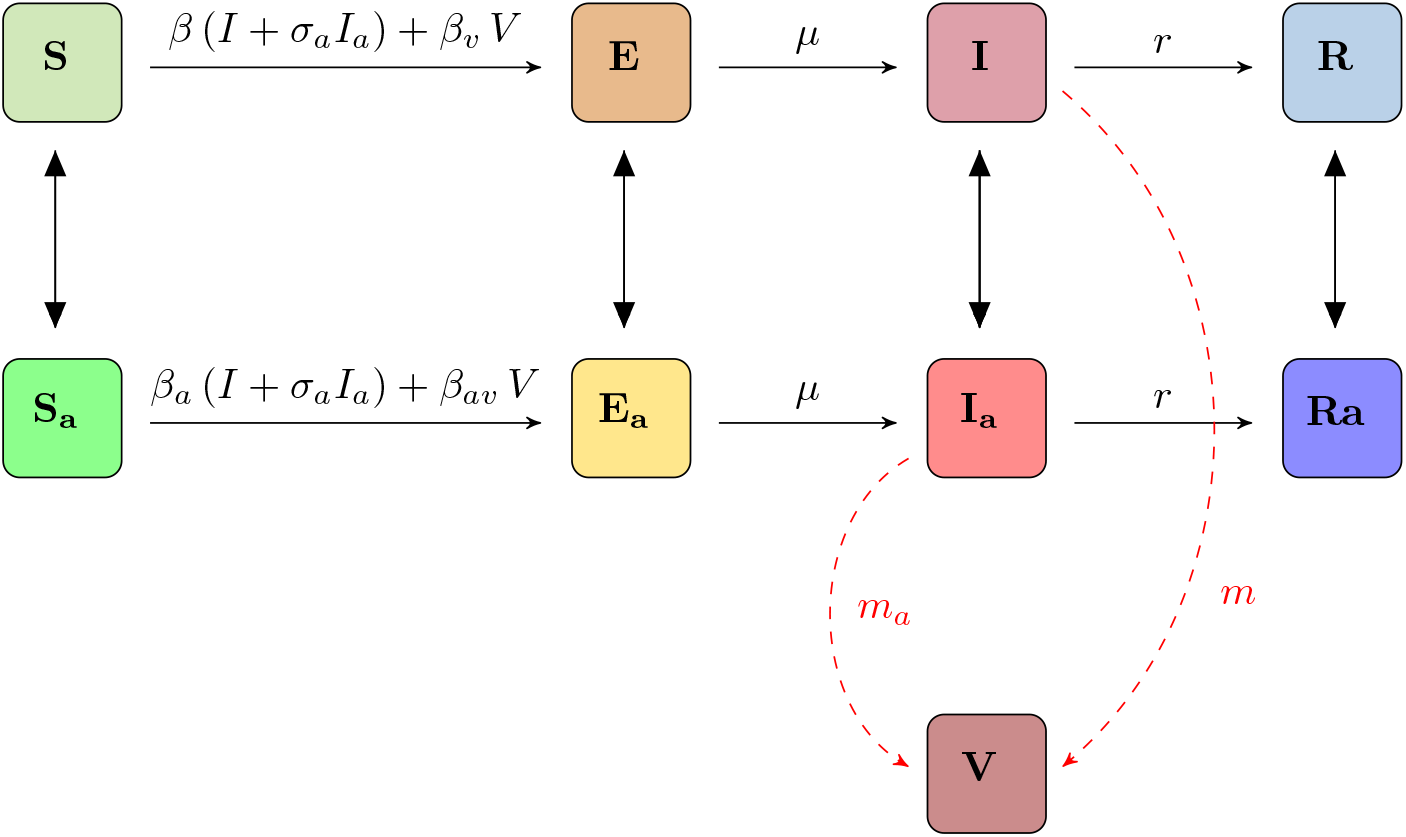
Schematic of the SEIR model. *Compartments for non-adherent population are as follows: Susceptible (S); exposed (E); Infected (I); and recovered (R). Corresponding compartments for adherent population are with subscript a. Individuals move from non-adherence to adherence at a rate γ and vice versa at a rate γ*_*a*_. *Black solid arrows show the flow of individuals between the compartments at rates indicated beside the arrows, while the dashed red arrows show virus shedding or contamination of surfaces by infected individuals (see* (2.0.1) *for more details)*.

## 3 Model analysis

In this section, we calculate the control reproduction number and the final size relation for the epidemic using the SEIRV model (2.0.1). We also consider some limiting scenarios of the epidemic and derive the control reproduction number and final size relations for these scenarios from the general control reproduction number and final size relation, respectively.

### 3.1 control reproduction number

We compute the control reproduction number for our model (2.0.1) using the next generational matrix approach of [21, 51] as used in [17]. To construct the Jacobian matrix of new infections *F* and that of transfer of infections *V* evaluated at the disease free equilibrium (DFE), we consider the exposed and infectious compartments of the SEIRV model (2.0.1) given by

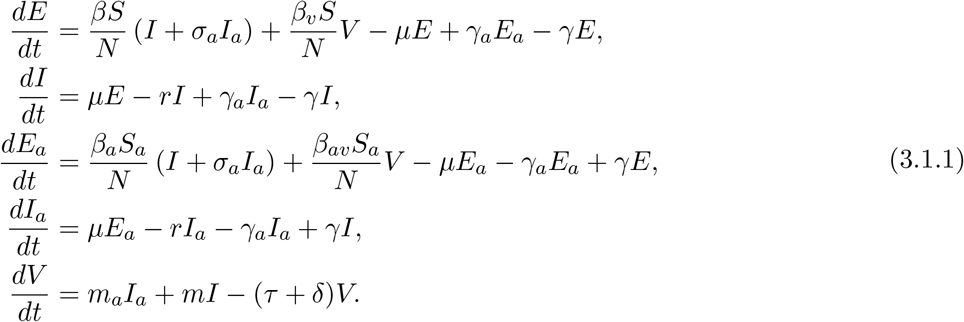

Upon using the relations between the transmission rates for the adherent and non-adherent populations given in (2.0.3), (3.1.1) reduce to

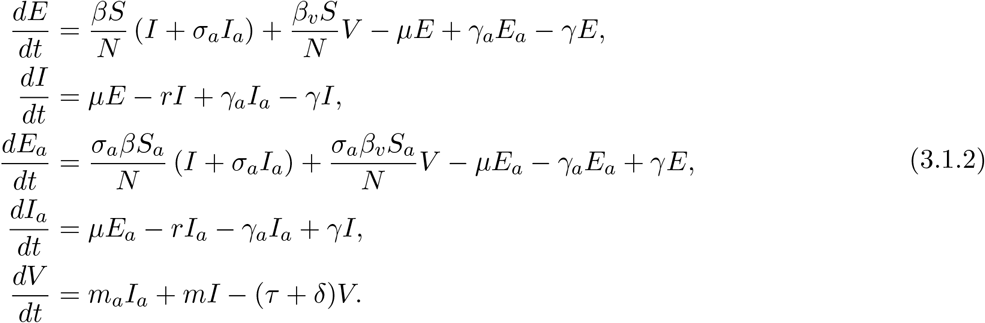

Recall that the adherence fraction in the population is given by *f* = *γ/*(*γ* + *γ*_*a*_). Therefore, at the beginning of the epidemic, *S*(0) = (1 − *f*)*N* = *N*_⋆_(0) and *S*_*a*_(0) = *fN* = *N*_*a*_(0). Following the arrangement of the equations in (3.1.2) and the variables *ϕ* = (*E, I, E*_*a*_, *I*_*a*_, *V*), at the DFE = (*N*_⋆_(0), 0, 0, 0, *N*_*a*_(0), 0, 0, 0, 0), the matrix for new infections *F* is given by

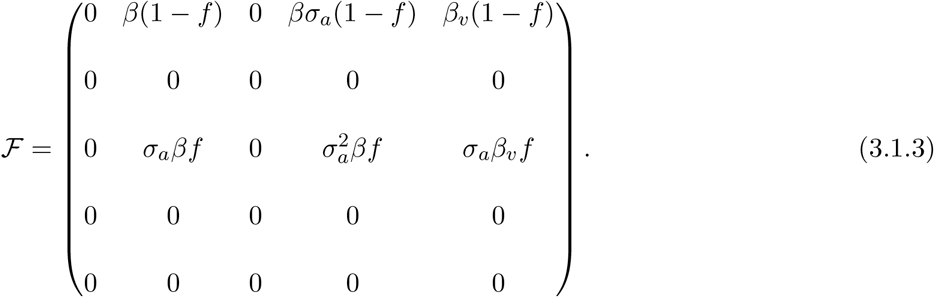

Similarly, the Jacobian matrix for transfer of infection at the beginning of the epidemic is given by

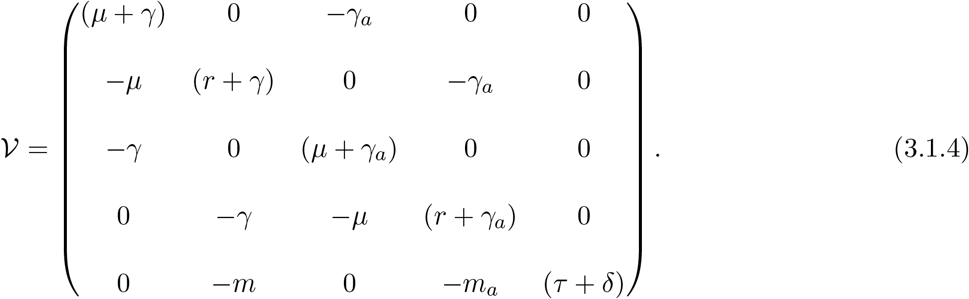

Upon computing the spectral radius of the next-generation matrix, ℱ𝒱^−1^ using the symbolic computational software SageMath [47], we obtain that the control reproduction number satisfies

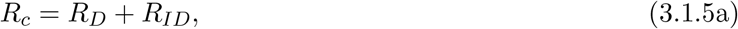

where *R*_*D*_, which accounts for disease transmission through direct route among non-adherent and adherent individuals is given by

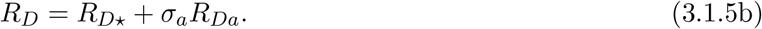

Here, *R*_*D*⋆_ and *R*_*Da*_ are defined as

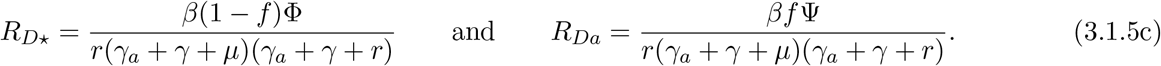

The second term in the control reproduction number (3.1.5a) accounts for indirect transmissions, and it is given by

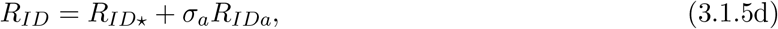

where *R*_*ID*⋆_ and *R*_*IDa*_ are the indirect transmissions contributed by the non-adherent and adherent populations, respectively, and are given by

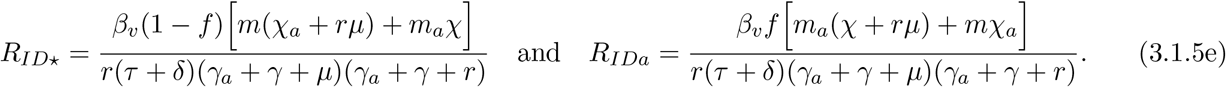

The newly introduced variables *𝒳, 𝒳*_*a*_, Φ, and Ψ are defined as follows:

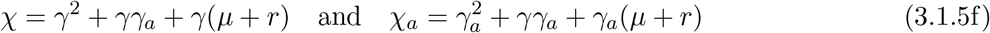

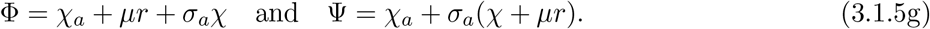

Suppose there is no adherent population, that is, *f* = 0, *σ*_*a*_ = 0, *γ*_*a*_ = 0 and *γ* = 0, we have Φ = *µr* and Ψ = 0. Therefore, (3.1.5a) reduces to

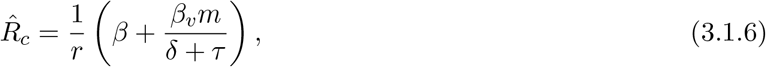

where 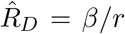 accounts for infections cause through direct transmission, while 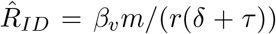 accounts from those of indirect transmission. Observe that 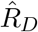 will increase as both the transmission rate *β* and recovery rate 1*/r* increase. This shows that an infected individual will spread the virus/disease more if the transmission rate of the disease is increases and if it takes longer for them to recover from the disease. For 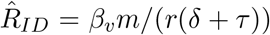, infections increases as the shedding/contamination rate of infected individuals increase and decrease as the virus decay rate *δ* and the environment cleaning or sanitization rate *τ* increases. Assuming a fraction 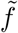 of the population were already adhering to COVID-19 NPIs before the disease was introduced into the population (as a precaution before the first case arrives) and there is no movement from adherent to non-adherent and vice versa, that is, *γ* = 0 and *γ*_*a*_ = 0, but *σ*_*a*_ ≠ 0, (3.1.5a) reduces to

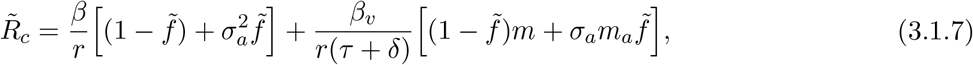

where the parameter *σ*_*a*_ with 0 ≤ *σ*_*a*_ ≤ 1 account for the reduction in acquiring or transmitting the disease due to adherence to COVID-19 NPIs. Similar to (3.1.5a) and (3.1.6), the control reproduction number 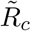 for this scenario also accounts for the contribution from both direct and indirect transmissions. Here, the contribution to 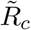 due to direct transmissions is given by

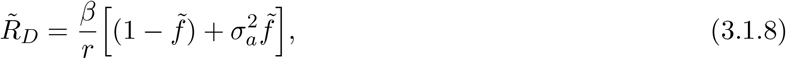

where 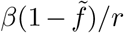 accounts for infections through direct transmissions caused by those that do not adhere to the NPIs, while 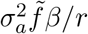 is the contribution from those that adhere to the NPIs. As one would expect, the transmissions caused by the adherent population will always be lesser than the infections caused by those that do not adhere since 0 ≤ *σ*_*a*_ ≤ 1. For this scenario, the contribution from indirect transmission is given by

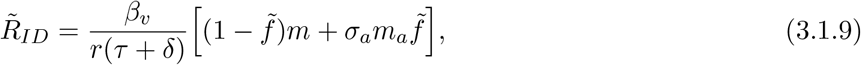

where 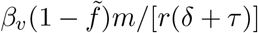 is the contribution from the non-adherent population and 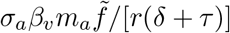 is from the adherent population. Note that 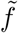 is the fraction of the population that adhere to the COVID-19 NPIs at the beginning of the epidemic.

### 3.2 Final size relation

Next, we derive a relation for the final epidemic size of our SEIRV model (2.0.1). This final size relation gives the total number of cases of the disease in the population during the epidemic. We consider a scenario where there is no movement between adherent and non-adherent susceptible populations. In other words, we assume that susceptible individuals do not change their behaviour from adhering to the COVID-19 NPIs to non-adherence and vice versa. This way, the equation for the susceptible populations *S* and *S*_*a*_ in (2.0.1a) and (2.0.1b), respectively, reduce to

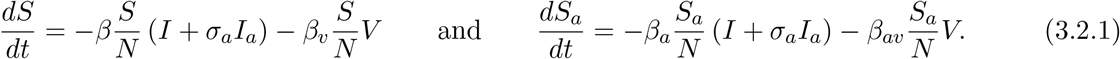

We assume that a fraction *f*, with 0 ≤ *f* ≤ 1, of the susceptible population adhere to COVID-19 NPIs before a disease outbreak, so that *S*(0) = (1 − *f*)*N* = *N*_⋆_(0) and *S*_*a*_(0) = *fN* = *N*_*a*_(0). In addition, we assume that a small number *I*(0) = *I*_0_ and *I*_*a*_(0) = *I*_*a*0_ are infected in the non-adherent and adherent populations, respectively, at the beginning of the epidemic and all other compartments are initially empty. Following the approach in [4, 6, 7, 8, 9, 10, 11, 12, 17, 18] and using the ODE system (2.0.1) together with the assumptions for the susceptible population in (3.2.1), we derive the final size relation for non-adherent population as

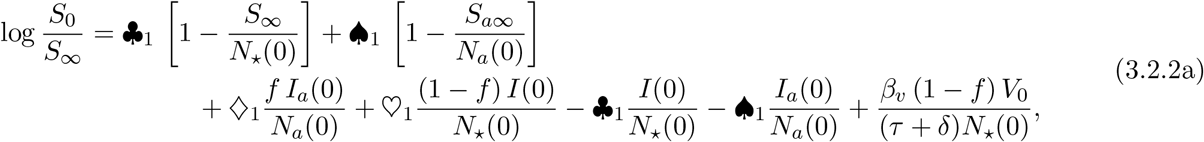

where ♣_1_ = *R*_*D*⋆_ + *R*_*ID*⋆_ and ♠_1_ = *R*_*Da*_ + *R*_*IDa*_ with *R*_*D*⋆_, *R*_*ID*⋆_, *R*_*Da*_ and *R*_*IDa*_ as given in (3.1.5). The remaining variables ♢_1_ and ♡_1_ are defined as follows

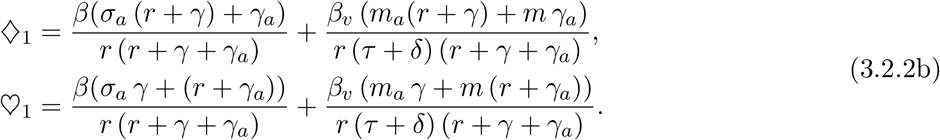

Similarly, the final size relation for adherent population is given by

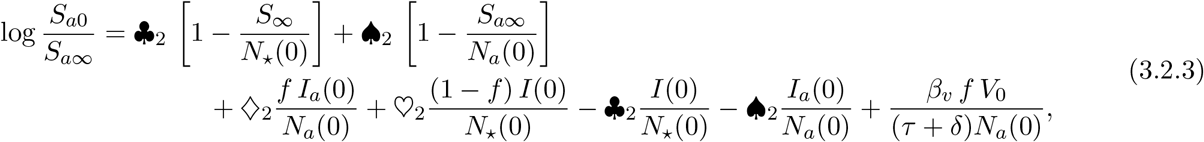

where ♣_2_ = *σ*_*a*_♣_1_, ♠_2_ = *σ*_*a*_♠_1_, ♢_2_ = *σ*_*a*_♢_1_ and ♡_2_ = *σ*_*a*_♡_1_ with ♣_1_, ♠_1_, ♢_1_ and ♡_1_ as given in (3.2.2). The final size relations in (3.2.2) and (3.2.3) for non-adherent and adherent population, imply that *S*_∞_ *>* 0 and *S*_*a*∞_ *>* 0, respectively. They give the relationship between the reproduction number *R*_*c*_ and the final epidemic size. The total infected population over the course of the epidemic is given by *N*_*a*_(0) − *S*_*a*∞_ and *N*_⋆_(0) − *S*_∞_ for the adIherent and non-adherent population, respectively, which can be described in terms of the attack rates as 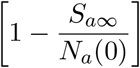 and 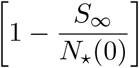 as in [7]. The final size relations in (3.2.2) and (3.2.3) can be written together in matrix form as

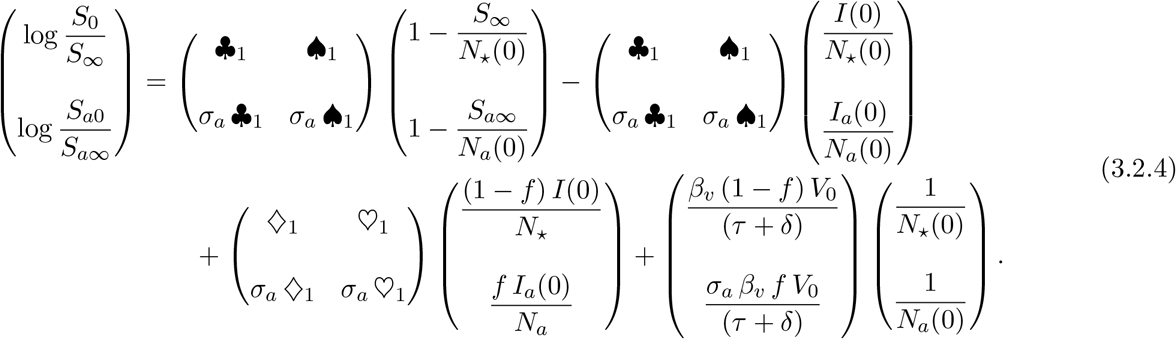

Also, from (3.2.2) and (3.2.3), we derive the following relation between the two final sizes

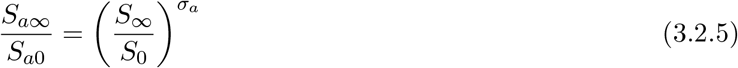

Since 0 ≤ *σ*_*a*_ ≤ 1, we have

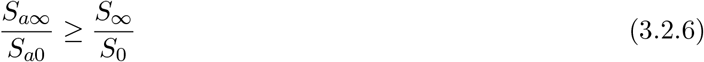

When 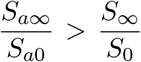, we conclude that the attack rate for the adherent population is smaller than that of the non-adherent population as a result of the reduction in the susceptibility and transmissibility of the adherent population. Observe that when *σ*_*a*_ = 1, which implies that there is no protection from adhering to NPIs, we have from (3.2.5) that *S*_*a*∞_*/S*_*a*0_ = *S*_∞_*/S*_0_. This implies that the attach rate is the same in both adherent and non-adherent populations, and as a result, we expect the epidemic size to be the same in both populations. This result agrees with our numerical simulation shown in red in the top left panel of Figure 4. On the other hand, when *σ*_*a*_ = 0, which implies that adhering to NPIs is 100% effective in preventing new infections, (3.2.5) reduces to *S*_*a*∞_ = *S*_*a*0_. This shows that there are no infections in the adherent population throughout the epidemic. Therefore, adherence to the NPIs during the pandemic is beneficial to reducing/containing the spread of the disease.

When the outbreak begins with no infected individuals, that is, the outbreak begins through an indirect transmission, the final size relations in (3.2.2) and (3.2.3) reduce to

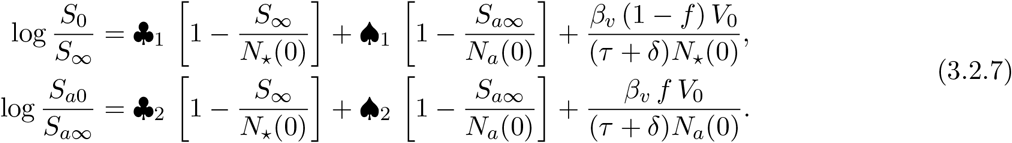

**Table 2:** Model parameters, descriptions, and values. *The direct and indirect transmission rates (β*_*a*_ *and β*_*av*_*), and shedding rate m*_*a*_ *for adherent population were derived from those of non-adherent population, respectively, using the relations in* (2.0.3).

| Parameter | Description | Value | Reference |
| --- | --- | --- | --- |
| $N$ | Total population | 5,000,000 | Assumed |
| $\sigma_a$ | Reduction in virus shedding, susceptibility, and onward transmission of disease due to adherence to NPIs | Varied | |
| $\beta$ | Direct transmission rate for non-adherence population | 0.5 | [3, 19, 39] |
| $\beta_v$ | Indirect transmission rate for non-adherence population | 0.15 | [19, 40] |
| $\beta_a$ | Direct transmission rate for adherence population | $\sigma_a\beta$ | Derived |
| $\beta_{av}$ | Indirect transmission rate for adherence population | $\sigma_a\beta_v$ | Derived |
| $\gamma$ | Rate of movement from non-adherent to adherent | 0.3 | [3, 16, 29, 31] |
| $\gamma_a$ | Rate of movement from adherent to non-adherent | 0.25 | [16, 29, 31] |
| $m$ | Virus shedding rate or the rate of contamination of surfaces by infected individuals in non-adherent population | 0.125 ml<sup>-1</sup><br>person<sup>-1</sup><br>day<sup>-1</sup> | [19, 34, 36] |
| $m_a$ | Virus shedding rate or the rate of contamination of surfaces by infected individuals in the adherent population | $\sigma_a m$ | Derived |
| $\delta$ | Decay rate of virus in the environment | 0.4 day<sup>-1</sup> | [14, 20, 59, 61, 62] |
| $\tau$ | Environment cleaning/sanitization rate | 0.1 | Inferred |
| $r$ | Recovery rate ($1/r$ is the infectious period) | 1/5 day<sup>-1</sup> | [3, 22, 27, 46, 48, 60] |
| $\mu$ | Rate of transitioning from exposed to infected ($1/\mu$ is the incubation period) | 1/6 day<sup>-1</sup> | [27, 35, 37, 60] |

Similarly, when the outbreak begins with only direct transmission with an infective, (3.2.2) and (3.2.3) become

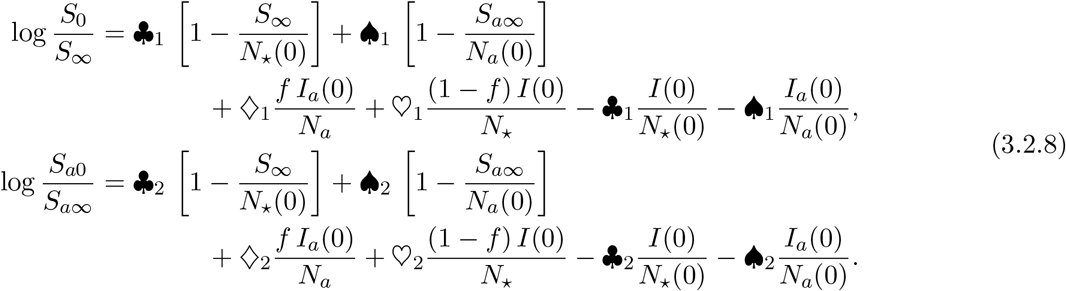

Now, consider an instance where there is no adherent population, so that *f* = 0, *σ*_*a*_ = 0, *γ* = 0, and *γ*_*a*_ = 0. For this scenario, we have 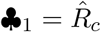, where 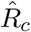 is as given in (3.1.6), ♠_1_ = 0, *I*_*a*_(0) = 0, *N*_⋆_(0) = *N*, ♣_1_ = ♡_1_. Also, since *σ*_*a*_ = 0, we have ♣_2_ = ♠_2_ = ♢_2_ = ♡_2_ = 0. This way, (3.2.2) and (3.2.3) reduces to

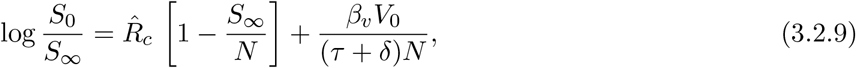

where 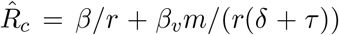 is the control reproduction number, given in (3.1.6), for the scenario without adherent population. As expected, when there is no adherent population, the final size relation reduces to that of a single population, and depends on the control reproduction number. When a fraction 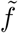 of the population adhere to COVID-19 NPIs but there is no movement between the adherent and non-adherent populations throughout the epidemic, that is, *γ* = *γ*_*a*_ = 0 and *σ* ≠ 0. The final size relation for non-adherent population given in (3.2.2) reduces to

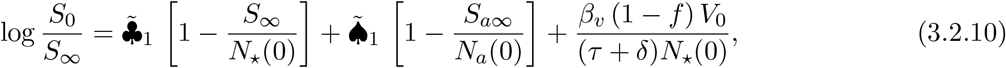

where

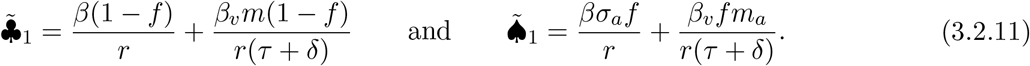

Similarly, for the adherent population, the final size relation (3.2.3) reduces to

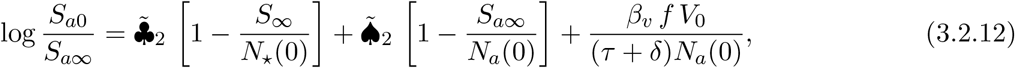

where 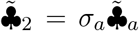 and 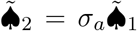 with 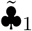 and 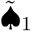 as defined in (3.2.11). Let 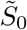 and 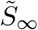 be the total susceptible population at the beginning and the end of the epidemic, respectively. Then at the beginning of the epidemic, we have

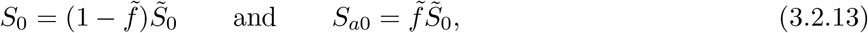

where *S*_0_ and *S*_*a*0_ are the initial susceptibles in non-adherent and adherent populations, respectively. Since there is no movement between the adherent and non-adherent population, the following conditions are satisfied at the end of the epidemic

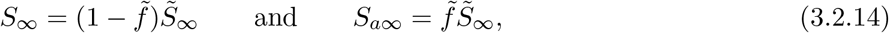

where *S*_∞_ and *S*_*a*∞_ are the susceptibles left in non-adherent and adherent populations, respectively, at the end of the epidemic. Upon adding (3.2.10) and (3.2.12), and using (3.2.13) and (3.2.14), and *m*_*a*_ = *σ*_*a*_*m*, we have

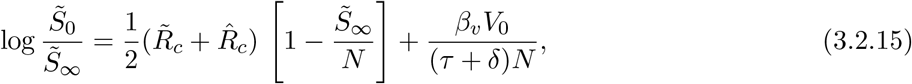

where 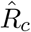 and 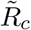are as given in (3.1.6) and (3.1.7), respectively.

In the next section, we present numerical computations of the control reproduction number (3.1.5) for different parameters of the model. We also present numerical simulations of the SEIR model (2.0.1), where we numerically computed final epidemic sizes for different scenarios.

## 4 Numerical simulations

In this section, we study the effect of different parameters of the model on the control reproduction number *R*_*c*_, given in (3.1.5). These results are presented as contour plots. In addition, we present numerical simulations of the SEIRV model (2.0.1) and investigate the effect of different parameters of the model on the disease dynamics and final epidemic size.

Table 2 shows the parameters of the SEIRV model (2.0.1) with their descriptions and values. Some of these parameters were obtained from published articles, while others were either inferred, derived or varied. References are provided for parameters that were obtained from published articles. The direct and indirect transmission rates (*β*_*a*_ and *β*_*av*_), and shedding rate *m*_*a*_ for adherent population were derived from those of non-adherent population, respectively, using the relations in (2.0.3).

In Figure 2, we present contour plots of the control reproduction number (3.1.5) computed as a function of different parameters of the model (2.0.1). The top left panel of this figure shows *R*_*c*_ as a function of the direct and indirect transmission rates. As one would expect, *R*_*c*_ increases as both transmission rates increase. We also notice that the direct transmission rate *β* has more effect on the control reproduction number than the indirect transmission rate *β*_*v*_. This may be because indirect transmissions are mitigated by environment cleaning and virus decay rate. In the top right panel of the same figure, we have *R*_*c*_ as a function of the adherence rate *γ* and the direct transmission rate *β*. This result shows that the adherence rate has little effect on the control reproduction number when the transmission rate is low, but has more effect on it as the transmission rate increases. When the transmission rate is low, there is a lesser chance of getting infected, as a result, adherence to the NPIs will have little effect on the spread of infection. However, when transmission rate is high, adhering to the NPIs will significantly reduce disease transmission. Similar result is shown in the middle left panel of Figure 2 for the adherence rate *γ* and the indirect transmission rate *β*_*v*_. For this scenario, *R*_*c*_ increases as the adherence to NPIs decreases even for a low indirect transmission rate, unlike the case of direct transmission (top right panel of Figure 2), where adherence rate does not affect *R*_*c*_ when *β* is small. The reason for this is that indirect transmission does not only depend on the transmission rate. It also depends on the environment cleaning/sanitization rate and the shedding and decay rate of the virus. If a lot of shedding is happening, or the decay rate of the virus is low, there may be a need to adhere more to the NPIs to reduce the spread of the disease.

**Figure 2:**
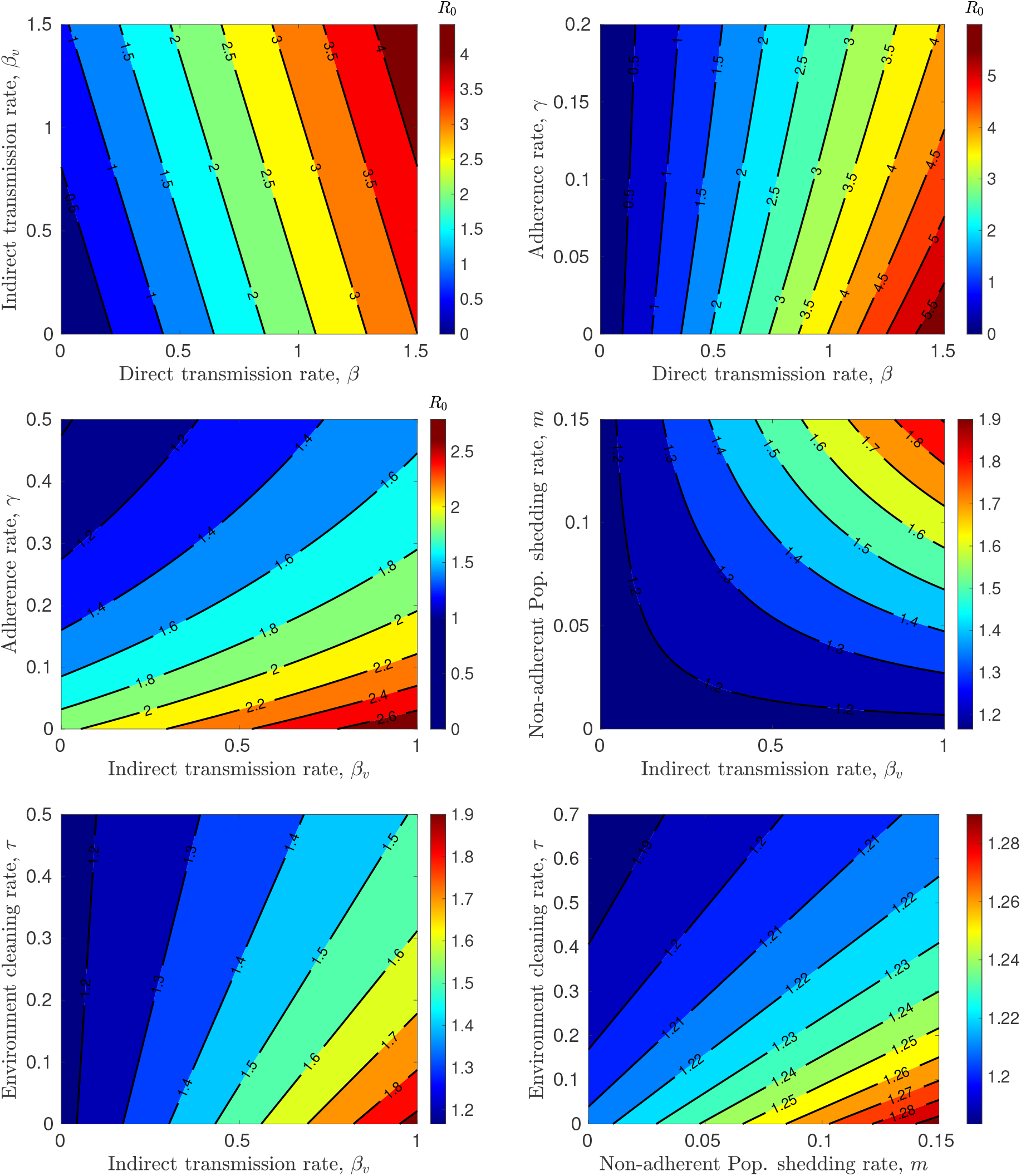
Computed control reproduction number. *Contour plots of the control reproduction number* (3.1.5) *computed as a function of different parameters of the model. Top left: the direct transmission rate β and indirect transmission rate β*_*v*_. *Top right: the adherence rate γ and direct transmission rate β. Middle left: the adherence rate γ and indirect transmission rate β*_*v*_. *Middle right: the shedding rate for non-adherence individuals m and indirect transmission rate β*_*v*_. *Bottom left: the environment cleaning rate τ and indirect transmission rate β*_*v*_. *Bottom right: the environment cleaning rate τ and the shedding rate for non-adherence individuals m. Parameters are as given in Table 2, except when used to generate the plot*.

For a fixed indirect transmission rate, *R*_*c*_ increases as the virus shedding rate increases, although, if the transmission rate is low, *R*_*c*_ remains constant irrespective of the shedding rate. Similarly, for a fixed shedding rate *m* for non-adherent population, *R*_*c*_ increases as *β*_*v*_ increases, but remains constant when shedding rate is too low irrespective of the value of *β*_*v*_ (see the middle right panel of Figure 2). This result shows that when infected individuals do not shed much virus or contaminate surfaces, there will be no indirect disease transmission through these surfaces. Similar result is obtained when *R*_*c*_ is computed as a function of the shedding rate *m*_*a*_ for the adherent population and the indirect transmission rate *β*_*v*_. In the bottom panel of Figure 2, we present *R*_*c*_ as a function of the environment cleaning/sanitization rate and the indirect transmission rate *β*_*v*_ (left panel), and the environment cleaning/sanitization rate and the shedding rate of non-adherent individuals *m* (right panel). Both results show that *R*_*c*_ decreases as the environment cleaning/sanitization rate increases, and increases with increase in the transmission and shedding *β*_*v*_ and *m*, respectively.

Figure 3 shows the total disease prevalence (total number of infected individuals at a given time) in the population over time for different rates of movement from adherent population to non-adherent and vice versa. These results are used to investigate the effect of adherence to COVID-19 NPIs on the dynamics of the disease. We consider four (4) different scenarios based on the rate of movement from adherent to non-adherent population *γ*_*a*_ and the rate of moving from non-adherent to adherent population *γ* for different values of the parameter *σ*_*a*_, used to model the reduction in susceptibility, shedding of virus, and onward disease transmission for the adherent population. The first scenario shown in the top left panel of Figure 3 is for *γ* = *γ*_*a*_ = 0, where there is no movement of individuals between non-adherence and adherence populations. For this scenario, the epidemic final size is 60.19% when *σ*_*a*_ = 0.5, 80.71% when *σ*_*a*_ = 0.75, and 91.43% when *σ*_*a*_ = 1. As expected, the largest epidemic occurs when there is no protection derived from adhering to the NPIs, that is, when *σ*_*a*_ = 1. Epidemic decreases as the protection from NPIs increases (*σ*_*a*_ decreases). Similar results are observed for the remaining scenarios in this figure. When there is movement between the adherent and non-adherent population such that the rate of going from adherence to non-adherence is the same as that of going from non-adherence to adherence, specifically, when *γ* = *γ*_*a*_ = 0.3, the final epidemic size is 59.50% for *σ*_*a*_ = 0.5, 81.20% for *σ*_*a*_ = 0.75 and 91.43% for *σ*_*a*_ = 1. The total prevalence plots for this scenario are shown in the top right panel of Figure 3.

**Figure 3:**
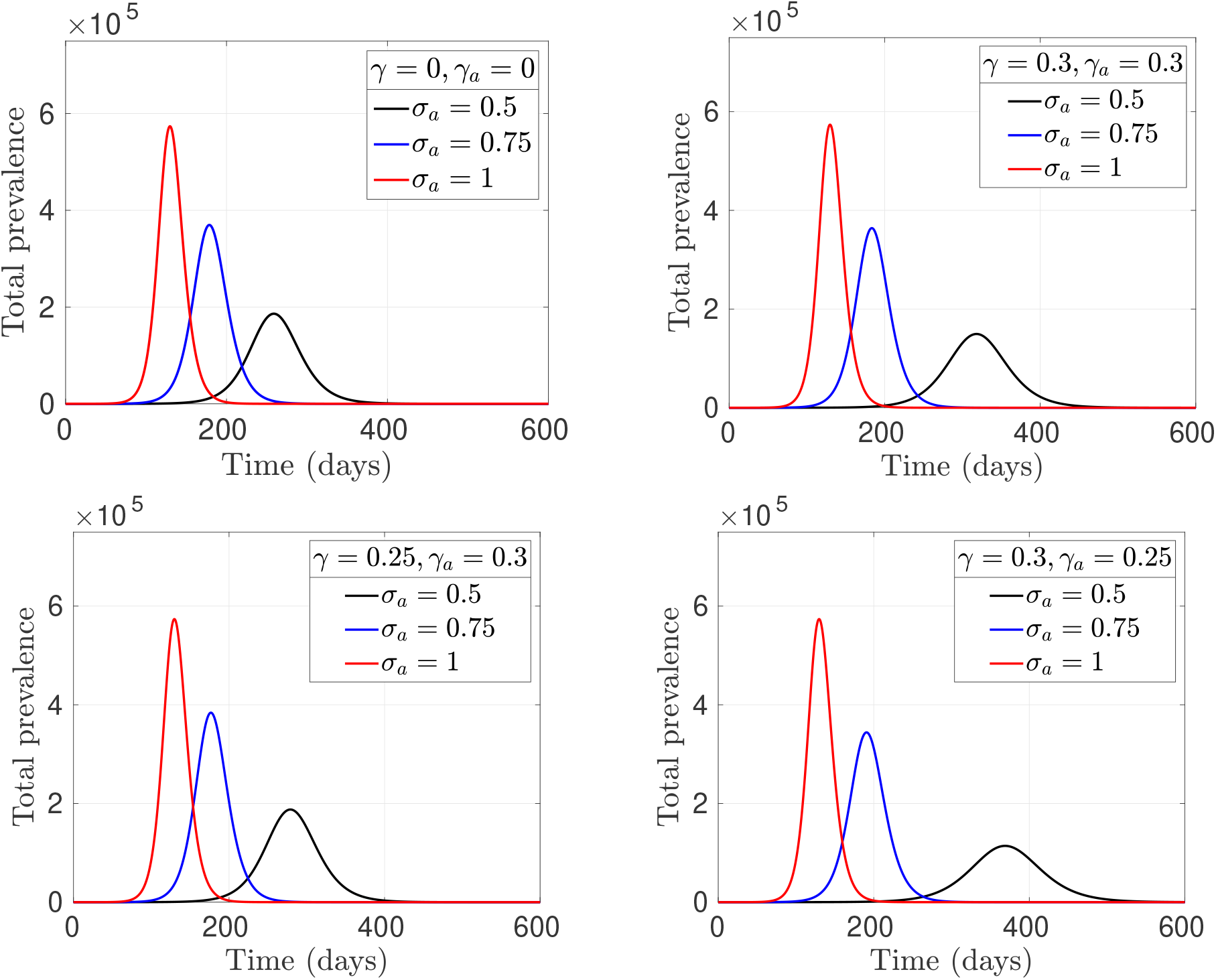
Effect of adherence to NPIs on COVID-19 epidemic. *The total disease prevalence over time for different values of the parameter σ*_*a*_ *(used to model reduction in virus shedding, susceptibility, and onward disease transmission for the adherent population) and the movement rates γ (non-adherence to adherence) and γ*_*a*_ *(adherence to non-adherence). Top left: γ* = *γ*_*a*_ = 0 *with* 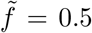, *top right panel: γ* = *γ*_*a*_ = 0.3, *bottom left: γ* = 0.25 *and γ*_*a*_ = 0.3, *and bottom right: γ* = 0.3 *and γ*_*a*_ = 0.25. *All other parameters are as given in Table 2 with initial conditions S*(0) = (1 − *f*)*N, E*(0) = 0, *I*(0) = 1, *R*(0) = 0, *S*_*a*_(0) = *fN, E*_*a*_(0) = 0, *I*_*a*_(0) = 1, *R*_*a*_(0) = 0 *and V* (0) = 1.

In the bottom panel of Figure 3, we show the results for *γ* = 0.25 and *γ*_*a*_ = 0.3 (left panel), and *γ* = 0.3 and *γ*_*a*_ = 0.25 (right panel). For the result in the left panel where the rate of movement from the adherent population to non-adherent (*γ*_*a*_) is higher than that of going from non-adherence to adherence (*γ*), the epidemic final sizes are 64.86%, 82.49% and 91.43% for *σ*_*a*_ = 0.5, *σ*_*a*_ = 0.75 and *σ*_*a*_ = 1, respectively. On the other hand, for *γ* = 0.3 and *γ*_*a*_ = 0.25 (bottom right panel of Figure 3), where the rate of movement from the non-adherence to adherence is higher than that of going from adherent population to non-adherent, the epidemic final size is 53.41% for *σ*_*a*_ = 0.5, 79.83% for *σ*_*a*_ = 0.75, and 91.43% for *σ*_*a*_ = 1. We observe from the results in this figure that the epidemic size decrease as *σ*_*a*_ decreases. This shows that fewer infections are occurring as people get more protection from adhering to the NPIs, since the benefit of adhering to NPIs increases as *σ*_*a*_ decreases. In addition, the epidemic size when *σ*_*a*_ = 1 (no protection due to adherence to NPIs) is the same for any values of *γ* and *γ*_*a*_. For *σ*_*a*_ = 0.5 and *σ*_*a*_ = 0.75, the smallest epidemics were predicted for the scenario where the rate of movement from non-adherence to adherence *γ* is higher than that of moving from adherence to non-adherence *γ*_*a*_ (bottom right panel of Figure 3). Whereas the largest epidemics occur when the rate of adherence *γ* = 0.25 is lower than that of non-adherence *γ*_*a*_ = 0.3. These results show that there will be smaller epidemic if more people adhere to the NPIs.

The results in Figure 4 were obtained using the same parameters and initial conditions as those in Figure 3 but plotted in terms of the adherent and non-adherent populations. Presenting the results in this way enables us to study the dynamics of the two populations as the movement rates *γ* and *γ*_*a*_ are varied. We notice from the results in this figure that even though the final epidemic size is the same for all four scenarios when *σ*_*a*_ = 1 (as discussed earlier in Figure 3), the dynamics of the disease is different for each scenario. When *γ* = *γ*_*a*_ are equal, the disease dynamics is the same for both adherent and non-adherent population when *σ*_*a*_ = 1 (see the red plots in the top panel of Figure 4). This is because the population is divided into two equal halves between the adherent and non-adherent population for these scenarios, and no protection is derived from adhering to the NPIs (since *σ*_*a*_ = 1). When *σ*_*a*_ ≠ 1, for the scenario with *γ* = *γ*_*a*_ = 0.3, the dynamics of the disease in the two populations are the same irrespective of *σ*_*a*_ (see top right panel of Figure 4). However, this is not the case for *γ* = *γ*_*a*_ = 0, even though *f* = 0.5 in both scenarios. For *σ*_*a*_ = 0.5 and *σ*_*a*_ = 0.75, there are more infections in non-adherent population than the adherent population. This is as a result of the protection derived from adhering to the COVID-19 NPIs. When *γ* = 0.25 and *γ*_*a*_ = 0.3, there are more infections in the non-adherent population than the adherent population for all values of *σ*_*a*_. This is due to the fact that there are more people in the non-adherent population since the non-adherence rate is higher (*γ*_*a*_ *> γ*) and they do not get protection from the NPIs. The most interesting and counter-intuitive result is when *γ* = 0.3 and *γ*_*a*_ = 0.25 (bottom right panel of Figure 4). This result shows that there are more infections in the adherent population than non-adherent population. We believe this is as a result of the adherent population size because more people are moving to the adherent population since *γ > γ*_*a*_. It is important to note that even though there are more infections in the adherent population than non-adherent population, the final epidemic size is still smaller for the *γ* = 0.3 and *γ*_*a*_ = 0.25 scenario, relative to that of the *γ* = 0.25 and *γ*_*a*_ = 0.3 scenario for *σ*_*a*_ = 0.75 and *σ*_*a*_ = 0.5, as discussed earlier and shown in the bottom panel of Figure 4. The epidemic size is the same for both scenario when *σ*_*a*_ = 1.

**Figure 4:**
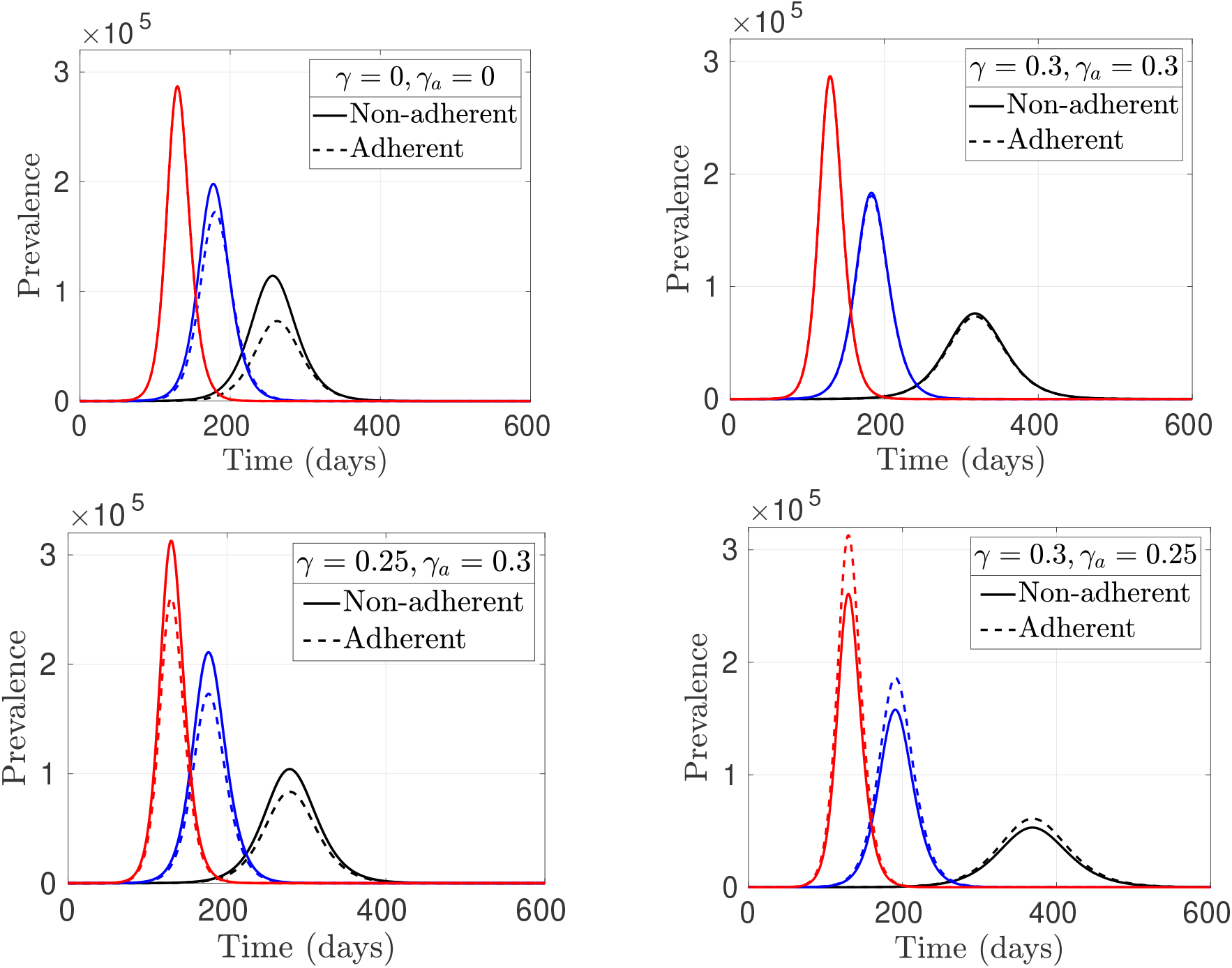
Dynamics of adherent and non-adherent populations. *Disease prevalence over time for adherent and non-adherent populations for σ*_*a*_ = 0.5 *(black), σ*_*a*_ = 0.75 *(blue) and σ*_*a*_ = 1 *(red). Top left: γ* = 0 *and γ*_*a*_ = 0 *with* 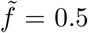, *top right panel: γ* = 0.3 *and γ*_*a*_ = 0.3, *bottom left: γ* = 0.25 *and γ*_*a*_ = 0.3, *and bottom right: γ* = 0.3 *and γ*_*a*_ = 0.25. *All other parameters are as given in Table 2 and initial conditions are the same as in Figure 3*.

In Figure 5, we investigate the effect of the indirect transmission rate *β*_*v*_ and the shedding rate *m* for non-adherent population on the dynamics of the disease. When the shedding rate is *m* = 0.02 (left panel of Figure 5), the final epidemic size is 46.66% for *β*_*v*_ = 0.15, 48.50% for *β*_*v*_ = 0.35, and 50.25% for *β*_*v*_ = 0.55. As the shedding rate increases to *m* = 0.125 (right panel of Figure 5), the final epidemic size for *β*_*v*_ = 0.15 is 53.44%, for *β*_*v*_ = 0.35 is 62.11%, and 68.87% for *β*_*v*_ = 0.55. This shows that the final epidemic size increases, as the transmission rate *β*_*v*_ increases. When the shedding rate is low, there is no much difference in the epidemic size as the transmission rate *β*_*v*_ increases. However, for a higher virus shedding rate or surface contamination rate, there is a significant difference in the final epidemic size as *β*_*v*_ increases. This is because when infected individuals do not shed much virus or contaminate shared surfaces, there will be no disease transmission even if the transmission rate (implicitly *R*_*c*_) is high. On the other hand, when a lot of shedding or surface contamination is happening, the disease will spread more easily leading to bigger epidemics as *β*_*v*_ increases. This results agree with the *R*_*c*_ computation shown in the middle left panel of Figure 2. A similar result was obtained for the shedding *m*_*a*_ for the adherent population (result not included).

**Figure 5:**
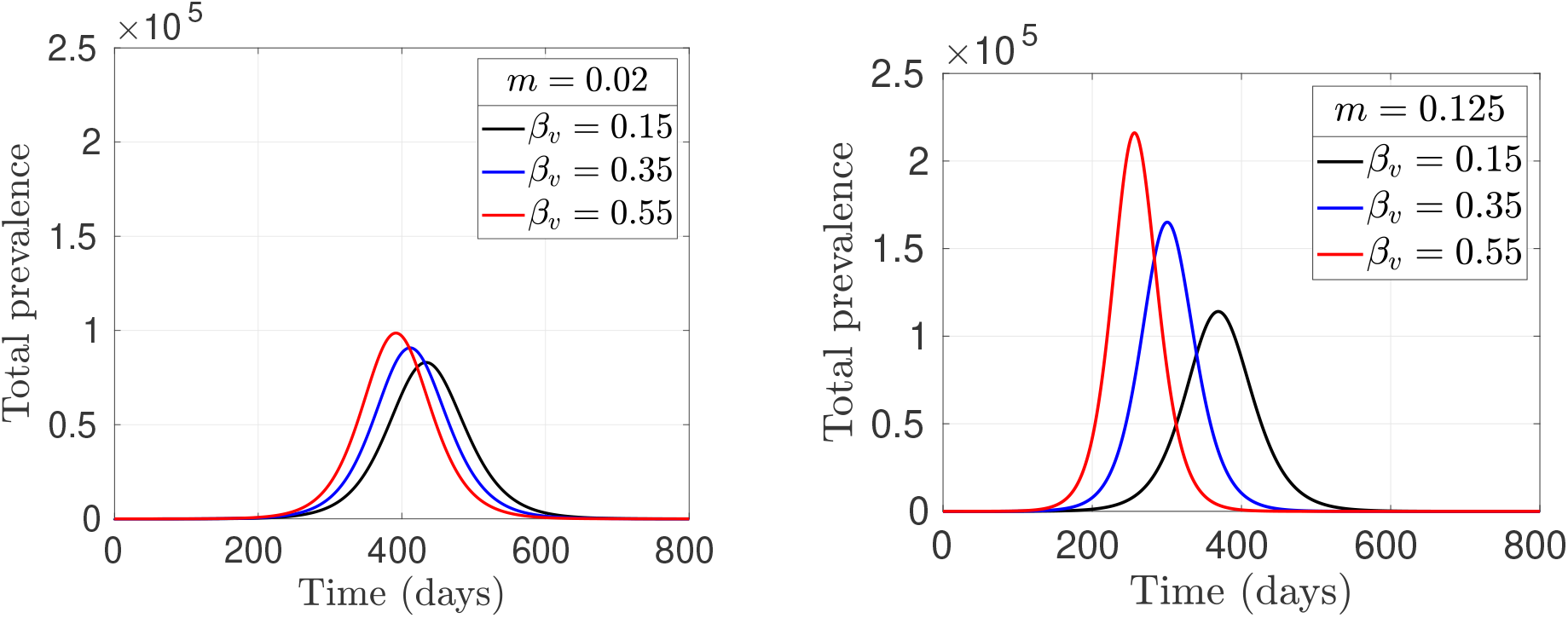
Effect of indirect transmission rate on disease dynamics. *Total disease prevalence over time for different values of the indirect transmission rate β*_*v*_ *and virus shedding (or surface contamination) rate m for non-adherent population. Left panel: m* = 0.02 *and right panel: m* = 0.125. *β*_*v*_ = 0.15 *(black), β*_*v*_ = 0.35 *(blue) and β*_*v*_ = 0.55 *(red). All other parameters are as given in Table 2 and initial conditions are the same as in Figure 3*.

Next, we study the effect of different initial conditions on the dynamics of the diseases. We consider two scenarios: when the epidemic starts through direct transmission and when it starts through indirect transmission. For the scenario where the epidemic starts through direct transmission, we assume that there are only two infected individuals in the population at the beginning of the epidemic; one infective in the adherent population and the other in the non-adherent population, that is, *I*_*a*_(0) = 1, *I*(0) = 1. And that there are no viruses in the environment *V* (0) = 0, while the remaining compartments are empty. On the other hand, when the epidemic starts with the indirect transmission, we assume that there are no infected individuals in the population at the beginning of the epidemic, that is, *I*_*a*_(0) = 0 and *I*(0) = 0, and *V* (0) = 1, with the remaining compartments empty. The results for both scenarios are shown in Figure 6 for different values of the movement parameters *γ* and *γ*_*a*_, where the solid curves are for the case where epidemic start through direct transmission, and the dashed curves are for when the epidemic starts through an indirect transmission. We observe from these results that for all the cases considered, the total prevalence increases more rapidly when the epidemic starts through direct transmission, compared to when it starts through an indirect transmission. However, the final epidemic size is the same irrespective of the initial conditions. This result agrees with the *R*_*c*_ computation in the top left panel of Figure 2 that shows that the control reproduction number is more sensitive to the direct transmission rate *β* compared to the indirect transmission rate *β*_*v*_.

**Figure 6:**
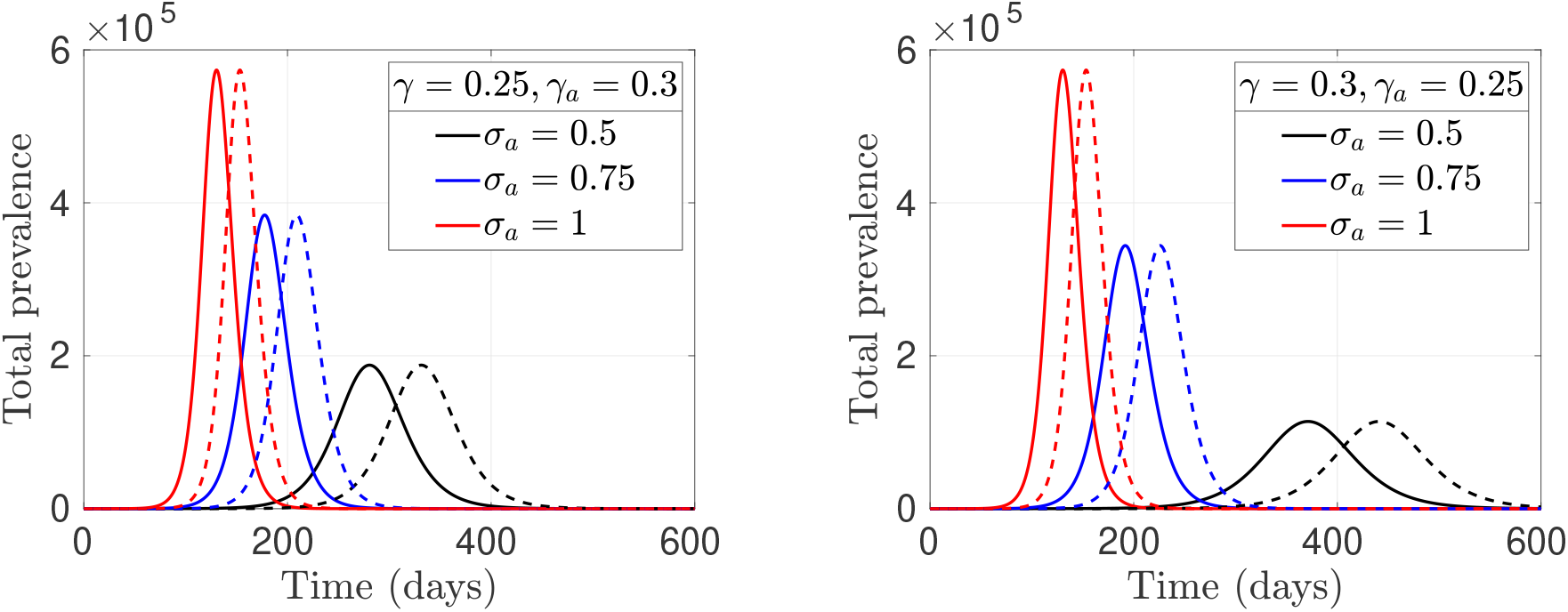
Effect of initial infection on disease dynamics. *Total disease prevalence over time showing the effect of different initial conditions on the dynamics of the diseases. The solid curves are for when the epidemic starts through a direct transmission with one infected individual in the adherent population and another in the non-adherent population, but no virus (I*_*a*_(0) = 1, *I*(0) = 1, *and V* (0) = 0*). The dashed curves are for when the epidemic starts through an indirect transmission with no infected individuals in both adherent and non-adherent populations (I*_*a*_(0) = 0 *and I*(0) = 0*) but V* (0) = 1. *Left panel: γ* = 0.25 *and γ*_*a*_ = 0.3 *and right panel: γ* = 0.3 *and γ*_*a*_ = 0.25. *σ*_*a*_ = 0.5 *(black), σ*_*a*_ = 0.75 *(blue) and σ*_*a*_ = 1 *(red). All other parameters are as given in Table 2*.

The results in Figure 7 show the effect of the environment cleaning or sanitization rate on the disease dynamics. When *β*_*v*_ = 0.15, the final epidemic size for *τ* = 0.1 is 53.44%, 51.26% for *τ* = 0.3, and 50.00% when *τ* = 0.5. As the indirect transmission rate increases to *β*_*v*_ = 0.5, the final epidemic size also increases with 67.33% for *τ* = 0.1, 62.38% for *τ* = 0.3, and 59.23% for *τ* = 0.5. Overall, the final epidemic size decreases as the rate of cleaning/sanitizing the environment increases. When the indirect transmission rate *β*_*v*_ is small, there is no much difference in the final epidemic size as *τ* increases, but for a higher transmission rate *β*_*v*_, an increase in the environment sanitizing rate significantly decreases the epidemic final size. These results are consistent with the *R*_*c*_ computation in the bottom left panel of Figure 2, where there is no significant difference in the control reproduction number as *τ* increase for a small value of *β*_*v*_, and a significant decrease in *R*_*c*_ as *τ* increases for a higher indirect transmission rate.

**Figure 7:**
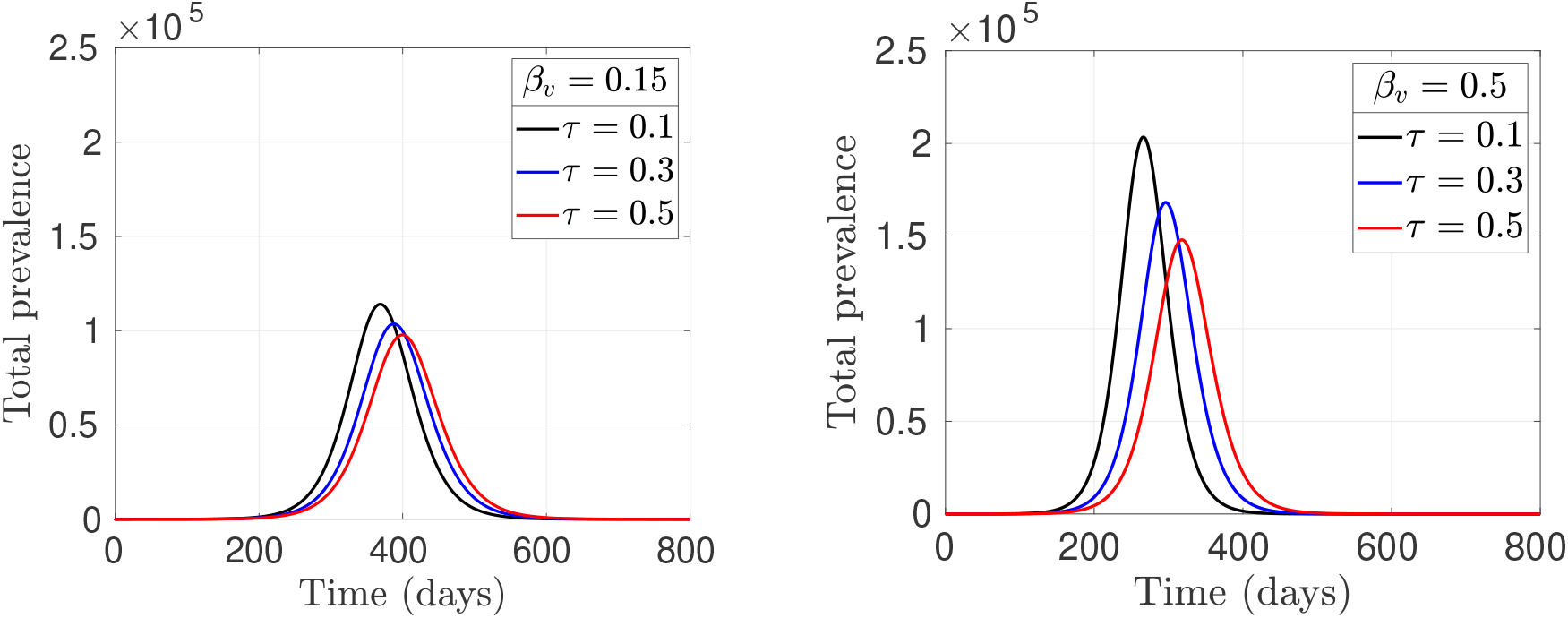
Effect of environment cleaning/sanitization on disease dynamics. *Total disease prevalence over time for different environment cleaning/sanitization rate τ and the indirect transmission rate β*_*v*_ *for non-adherent population. Left panel: β*_*v*_ = 0.15 *and right panel: β*_*v*_ = 0.5. *τ* = 0.1 *(black), τ* = 0.3 *(blue), and τ* = 0.5 *(red). All other parameters are as given in Table 2 and initial conditions are the same as in Figure 3*.

## 5 Discussion

We have developed and analyzed a SEIRV model for studying the dynamics of COVID-19. Our model divides the population into two groups: adherent and non-adherent, and considers both direct and indirect transmission of the disease. Individuals in the adherent population are assumed to adhere to all COVID-19 non-pharmaceutical interventions (NPIs) such as hand washing, physical distancing, wearing face masks, and avoiding large gatherings. In contrast, those in the non-adherent population do not adhere to these NPIs. By adhering to the NPIs, those in the adherent population have reduced their susceptibility, transmissibility and infectiousness. By direct transmission, we mean disease transmission that occurs when a susceptible individual comes in contact with an infected individual, while indirect transmission refers to the infections that occur through contaminated shared surfaces and objects such as door handles, utensils, etc. Individuals can move from adherent to non-adherent and vice versa at some rate in each human compartments.

We calculated the control reproduction number and final size relation for our model and studied the effect of the model parameters on the calculated control reproduction number. These results are presented as contour plots in Figure 2 and show that the direct transmission rate has more effect on the control reproduction number than the indirect transmission rate. We believe this is because indirect transmission depends on other factors such as shedding rate and environment cleaning rate. Similarly, when we studied *R*_*c*_ with respect to the adherence rate *γ* and the direct transmission rate *β*, we notice that adherence to NPIs has little effect on *R*_*c*_ when the transmission rate is small. As the direct transmission rate increases, our results show that adhering to the NPIs would significantly reduce *R*_*c*_. On other hand, adherence to NPIs is beneficial irrespective of the values of the indirect transmission rate. We also showed that the virus shedding rate has little effect on the control reproduction number if the indirect transmission rate is too small. However, as the transmission rate increases, an increase in shedding rate increases the control reproduction number significantly.

Furthermore, we studied the effect of some model parameters on the disease dynamics. We showed that the disease dynamics is different when there is no movement between adherent and non-adherent population, compared to when the movement between the two groups happens at the same rate. When there is no movement between the groups, there are less infections in the adherent population compared to the non-adherent population as a result of protection from adhering to NPIs. On the contrary, when movement happens between the two groups at the same rate, the level of infection in the two populations is the same, simply because the entire population becomes homogeneous as a result of the movement between adherent and non-adherent population happening at the same rate. When the non-adherence rate is higher than the adherence rate, there are more infections in the non-adherent population relative to the adherent population. In a situation where the adherence rate is higher than the non-adherence rate, the infection in the adherent population is higher compared to the non-adherent population, although, one would have expected the infections in the adherent population to be low because of the protection from adhering to the NPIs. This counterintuitive observation could be as a result of several factors, some of which includes our choice of movement parameters *γ* and *γ*_*a*_, and the parameter *σ*_*a*_ used to model protection in the adherent population due to NPIs. As the protection derived from adhering to the NPIs increases, fewer adherent individuals are getting infected. Overall, even though there are more infections in the adherent population when the rate of movement from non-adherent to adherent is higher than that of moving vice versa, the final epidemic size is smaller for this scenario than the other scenario. This result shows the benefits of adhering to the COVID-19 NPIs in reducing the epidemic size.

Also, we examined the effect of different initial conditions on the dynamics of the diseases. We considered when the epidemic starts through direct transmission and when it starts through indirect transmission. For the scenario where the epidemic starts through direct transmission, we assume that only two infected individuals are in the population at the beginning of the epidemic; one individual in the adherent population and the other in the non-adherent population (no contaminated surfaces), while the remaining compartments are empty. On the other hand, when the epidemic starts through indirect transmission, we assume that there are no infected individuals in the population at the beginning of the epidemic. However, there are some viruses on contaminated surfaces from which the first infections start. Our results show that the epidemic increases more rapidly when it starts through direct transmission, compared to when it starts through an indirect transmission, although the final epidemic size is the same irrespective of the initial conditions. Finally, we study the effect of environment sanitization on disease dynamics. Our results show that environmental sanitization reduces the prevalence of COVID-19 and consequently reduces the final epidemic size.

An immediate and interesting future direction in this project is to fit the model to reported cases of COVID-19 in a specific location and use region-specific parameters. By fitting the model to the cases data, important parameters that are not immediately available can be estimated from the data. The model can then be used to make forecasts/predictions and test the effect of implementing different intervention strategies. Another important future direction is to include vaccination compartments into the model. The COVID-19 vaccines became available and have been administered since early 2021. As of July 6, 2021, over 2.9 billion doses of the vaccine have been administered [53]. To properly capture the dynamics of COVID-19 in any population starting from January 2021, vaccination needs to be included in the model. It would be worthwhile to stratify the population by age so that the model can be used to answer age related questions such as those of school reopening and age-specific vaccination plans. There are currently four SARS-CoV-2 variants of concern (VOC) that are believed to be in circulation worldwide. These VOC are believed to be more contagious, leading to more severe sickness than the initial SARS-CoV-2 virus [56]. An interesting extension of our model would be to incorporate these variants of concerns.

Our model is prone to some limitations. This includes our assumptions that the population is well-mixed. We know contact rates vary from one person to another depending on their age group and activity level. Also, mixing patterns are different for individuals of different age groups. Using homogeneous mixing, we assume that everyone in the population has the same contact rate and mixing pattern. Another limitation of our model is that we have clustered all COVID-19 non-pharmaceutical interventions (NPIs) into one group, assuming that any individual that adheres to one of them will adhere to all. This may not be the case in reality. Some individuals may adhere to only a few of the NPIs. It would be nice to distinguish between the NPIs and study the effect of different NPIs on the disease dynamics. Similarly, we have assumed that the rate of movement from adherence to non-adherence and vice versa are constant over time. In a real-world scenario, these rates may change from time to time and may be affected by government policies and their implementation of the NPIs. Despite these limitations, our model can capture the overall dynamics of the COVID-19 epidemic considering infections transmitted through direct and indirect routes, and with and without adherence to the COVID-19 NPIs.

## Data Availability

N/A

## Acknowledgement

This work was conducted while JD was on maternity leave from a Postdoctoral position at York University.

